# Facial cleanliness, environmental factors and the burden of trachoma in a pastoral conflict area of Tiaty East Baringo County, Kenya

**DOI:** 10.1101/2025.10.07.25337475

**Authors:** Wanjira Njogu.L, George A. Makalliwa, Bridget Kimani, Kellen Karimi, Zephania Kibet, Doris Njomo, Titus Watitu, Paul M. Gichuki

## Abstract

Trachoma remains the leading infectious cause of preventable blindness globally, disproportionately affecting marginalized and impoverished communities. Despite global and national efforts guided by the World Health Organization’s (WHO) SAFE strategy (Surgery, Antibiotics, Facial cleanliness, Environmental improvement), the latter two components remain under-implemented. In Kenya, trachoma remains endemic in 13 of 47 counties, including Baringo.

A cross-sectional study was conducted in Tiaty East Sub-County, a trachoma Sub-Evaluation Unit in Baringo, to determine the burden of trachoma and assess facial cleanliness and environmental conditions. Using a structured questionnaire and observation checklist, data were collected from 178 randomly selected households. A total of 279 children aged 1–9 years were examined for trachomatous inflammation–follicular (TF).

The overall TF prevalence was 5.02%, slightly above the WHO elimination threshold of <5%. Facial cleanliness was sub-optimal with 91.8% of children having visibly unclean faces. Latrine coverage was low (21.3%), and 99.1% of those without latrines practiced open defecation. Most households (61.8%) sourced water from a dam, with 80.9% requiring more than 30 minutes to fetch water. Handwashing stations were absent in 94% of households. Multivariable analysis revealed caregiver age and awareness of poor hygiene as a risk factor for trachoma and were also significantly associated with TF.

The persistence of TF and poor F&E indicators highlight critical gaps in trachoma control in the Sub-Evaluation Unit. Strengthening implementation of the F&E components of the SAFE strategy is essential to achieving trachoma elimination in this setting.

**Author summary:** Trachoma is an eye disease that can cause blindness if not controlled. Kenya has made great progress towards eliminating trachoma by 2027, but challenges remain in pastoralist communities. Because these communities are often on the move, it is difficult to provide consistent health services and to improve water, sanitation, and hygiene. Facial cleanliness and environmental hygiene improvement are especially important for stopping the spread of infection and should be part of the larger intervention to eliminate trachoma. In our study, we found that some areas still have signs of infection and gaps in hygiene and environmental improvements. Addressing these gaps is essential if Kenya is to meet the World Health Organization’s target for trachoma elimination and restore dignity to the community by breaking the disease transmission.

## Introduction

Trachoma is currently the leading infectious form of blindness, accounting for approximately 1.9 million forms of blindness globally with 103 million people living in endemic areas(1). Despite the recent efforts to eliminate the disease, trachoma remains a significant public health challenge in most countries, with approximately 115.7 million people globally living in trachoma endemic regions(2). The World Health Organization (WHO) estimates that the annual economic burden attributed to loss of productivity through trachoma is $ 8 billion(1). The disease is considered endemic in 38 countries globally, where the countries are mostly located in Asia, Africa and the Middle East(1). Sub Saharan Africa bears a substantial burden of trachoma, as 59% of the countries are endemic and 85% of people live with trachoma(3)

The World Health Organization, in 1996, embarked on an audacious international program to control and preferably eliminate trachoma under a global program titled, “Global Elimination of Trachoma” (GET) by the end of 2020 where one of the targets was to reduce the prevalence of Trachoma Inflammation Folliculitis (TF) to <5% (4). As part of the program, the WHO implemented the SAFE strategy which is a four-pronged plan for trachoma elimination, which includes Surgery, Antibiotics, Facial Cleanliness, and Environmental improvement(5). This is a comprehensive strategy particularly recommended for trachoma endemic regions. There has been expansive work in the implementation of the SAFE strategy across the globe (4). Studies have highlighted the efficacy of the strategy, noting significant progress towards the elimination of trachoma. A pilot project rolled out in Kajiado, Kenya in 2003, found that antibiotics helped reduce TF prevalence from 46.4% to 16.0% (6). However, the SAFE strategy has been implemented to varying degrees of success (7).There has been a general emphasis on the treatment part, which entails surgery and administration of antibiotics (S&A), with notable inconsistencies in the implementation of the Facial Cleanliness and Environmental improvement (F&E) aspects (8,9).

Facial cleanliness is defined as the absence of nasal and ocular discharges on the face, since these secretions attract eye-seeking flies and facilitate transmission of *Chlamydia trachomatis*, the bacteria that causes trachoma (10). Environmental improvement includes factors such as the presence and proper use of latrines, accessible handwashing stations, improved housing conditions (e.g., uncrowded households, adequate separation from domestic animals), and good personal hygiene practices such as not sharing fomites and regularly washing clothes (10,11). These measures reduce the presence of transmission means (ocular/nasal secretions) and vectors (flies), thereby lowering the risk of infection(12). When well implemented, F&E interventions are essential for interrupting transmission and sustaining elimination following surgery and antibiotic efforts(13). However, the effectiveness of F&E depends heavily on water, sanitation and hygiene (WASH) infrastructure and behavioral change in endemic communities, which make them complex but vital components of trachoma control(10).

In Kenya, trachoma is endemic in 13 counties including Isiolo, Kajiado, Turkana, Laikipia, Samburu, Marsabit, West Pokot, Embu, Kitui, Narok, Meru, Elgeyo Marakwet and Baringo with an estimated 2.1 million people living in these areas demanding interventions(14). The country established the Kenya Trachoma Elimination Program (KTEP), which is domiciled within the Ministry of health (MOH) with an ambitious five-year goal to eliminate trachoma as a public health problem by 2019 (15) and now aims to eliminate the disease by 2027(16). In October 2011, the Ministry of Health and the Department of Health Services launched Mass Drug Administration (MDA) strategy for trachoma, particularly in Tiaty East and Tiaty West Sub-Counties of Baringo County which together form part of a trachoma Evaluation Unit(EU) and had a baseline TF prevalence of 34.3% (16). Five rounds of MDA were implemented up to 2017, and after an impact survey in 2018, the WHO TF threshold had not been achieved (12.8%) informing three more rounds of MDA between 2020 to 2023 (16). The impact survey done after the last round of MDA in 2023 showed that the TF prevalence in the EU had dropped (3.02%) therefore achieving the WHO threshold of <5%)(17). A surveillance survey is scheduled for the end of 2025 to confirm if the prevalence is still below the elimination threshold post halting of MDA in the area.

While Baringo County has seen a significant reduction of TF from 34.3% in 2011 to 3.02% in 2023 due to intensive MDA interventions, limited access to the resident population due to insecurity and nomadic lifestyle poses a challenge in effectively reaching members of the community through MDA(16). A large proportion of the land is arid and semi-arid with poor Water, Sanitation, and Hygiene(WASH) infrastructure (18). In 2019, only 39% of the households had access to safe drinking water (19). The county therefore presents unique conditions, which can exacerbate the prevalence of trachoma by hindering the effective administration of the SAFE strategy, particularly the F&E components of the strategy.

Children aged 1-9 years, who are the main reservoirs of the *Chlamydia trachomatis* bacteria (11), are highly dependent on parents and caregivers for their hygiene needs. However, the nomadic lifestyles combined with high rates of poverty and illiteracy (20) could predispose most children to poor hygiene. Besides the impact surveys, few studies have focused on the implementation of the F&E components of the SAFE strategy in the county. While F&E enhancement is more cost-effective and sustainable ways of preventing and eliminating trachoma among other infectious diseases, few studies have examined household F&E aspects in marginalized communities. Gaps in such context specific evidence could continue hindering adoption and long-term utilization of the F&E best practices (12). Baringo County offers a unique context with an intersection of factors that could potentially impact on the effectiveness of F&E as well as exacerbating the disease, hence a good context for the study. To this regard, our study examined the household prevalence of TF among children aged from 1-9 years in Tiaty East Baringo County and its association with household F&E components of the SAFE strategy.

## Methods

### Ethics statement

This study was approved by the Kenya Medical Research Institute (KEMRI) Graduate School after receiving an ethical clearance from the KEMRI Scientific and Ethics Review Unit (SERU No 4986). A research permit was granted by the National Commission for Science Technology and Innovation (NACOSTI) (NACOSTI/P/24/41122). The study further received approval from Baringo County and sub county offices. A written informed consent in the local language (Pokot) was obtained from heads of households or caregivers aged above 18 years. Names of the participants were anonymized by a unique identifier for confidentiality purposes.

### Study area

The study was conducted in Tiaty East Sub-County, Baringo County (*Figure 1*). Tiaty East covers an area of 2,106.5 km^2^ with a population of 73,146 and 12,153 households. It is one of the seven sub-counties of Baringo County, which is in the former Rift Valley region. The County is home to Lake Baringo and has a total of 110,649 households(16). Tiaty East natives are mainly pastoralists who migrate during dry season and come back to reunite with families during rainy seasons (16). The main language of communication in the area is Pokot and cattle trading including sheep, goat, cows, camels and donkeys, is their main economic activity. Tiaty East sub-county is also characterized by a wide range of conflicts due to cattle rustling(16).

**Figure 1.**
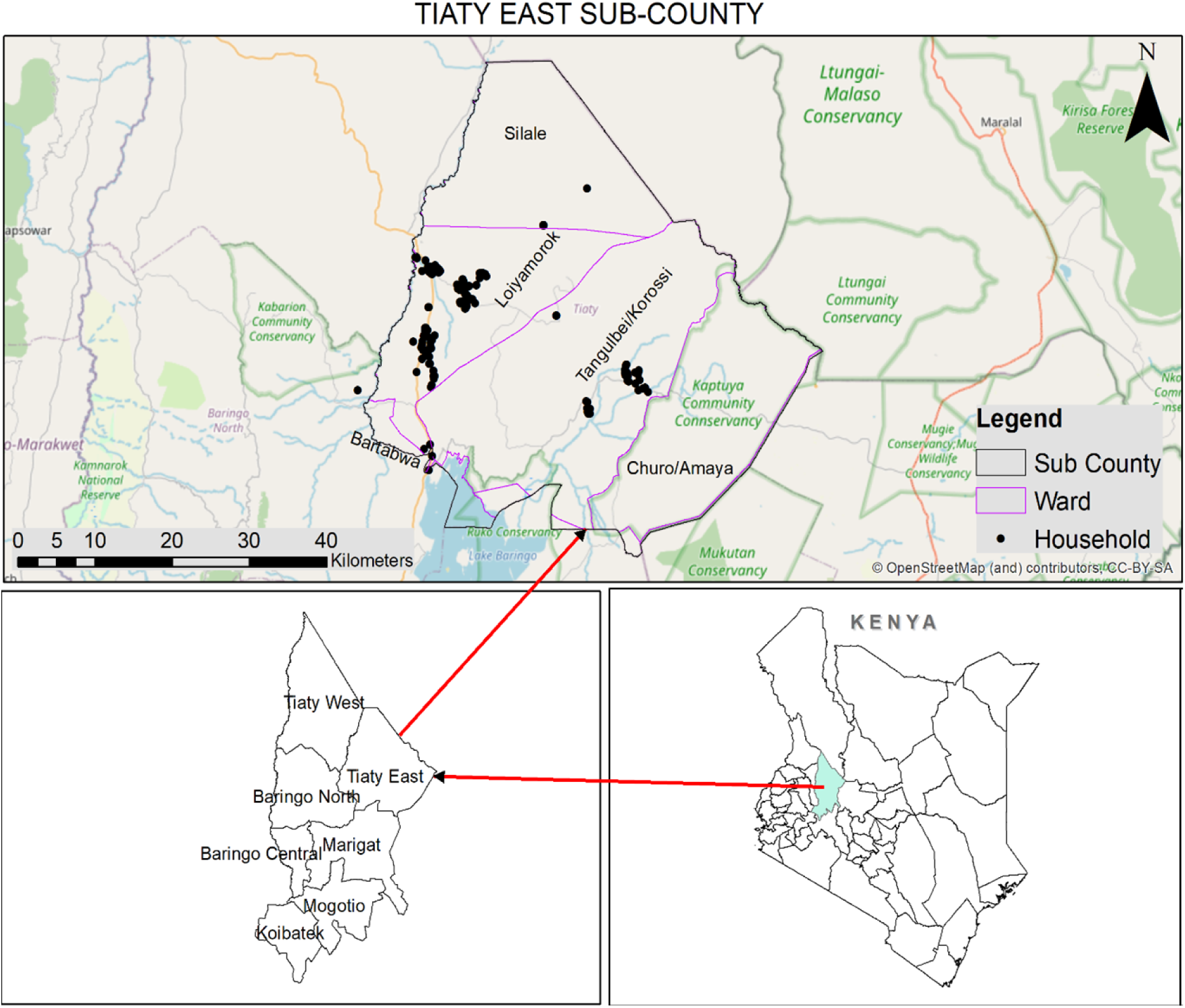
Map of study area showing sampled households (Geographic distribution of households captured during the study and created using ArcGIS 10.8. Basemap source: Esri, HERE, Garmin, FAO, NOAA, USGS, © OpenStreetMap contributors, and the GIS User Community.)

### Study design

This study used a cross-sectional research design, where data on household F&E factors and the prevalence of TF were collected at the same point in time. The identified positive cases were referred for treatment and follow up by Community Health Promoters (CHPs) who are links between the healthcare systems and the communities.

### Sample size calculation

The sample size was determined using Cochran’s formula with a prevalence of 5.9% (21), a design effect of 2.0 to adjust for clustering, and adjusted for 10% attrition. This led to a sample size of 190 households.

### Sampling and sample population

The study population comprised households in Tiaty East Sub-county, an area purposively selected due to its endemicity for trachoma and contextual challenges of pastoralism and conflict, which affect the implementation of the SAFE strategy (16). Within the sub-county, the sampling process followed several levels. A list of all administrative wards was obtained from the Sub-County Disease Surveillance Office. Three wards (Tangulbei, Silale, and Loiyamorok) were purposively selected to serve as strata, based on their population distribution and trachoma burden. Within each ward, villages were treated as clusters and randomly selected using simple random sampling. The villages selected were 18 in total: Achukut, Adadat, Atirirai, Chemakil, Chemoril, Cheptaran, Cherelio, Chesimirion, Dapalkow, Lokenoi, Lokrakow, Loruk, Manampelei, Oroo, Pilil, Riongo, Tunatai and Tuwo. In each selected village, households were sampled using a random walk method until the allocated sample size per cluster was achieved. The household served as the primary sampling unit. All eligible individuals within the sampled households were included in the survey, with the household as the main unit of analysis for statistical purposes. Households with children between 1 and 9 years old who had lived in the area for at least six months were included in the study.

### Data collection and management

A household pre-tested, structured questionnaire was administered to consenting households by trained research assistants. TF cases were assessed by the research assistants trained by an Ophthalmic Clinical Officer on eye-grading based on WHO criteria developed by Thylefors et al. (22), and cases confirmed by certified eye graders. An observational checklist was used to examine hygiene, environmental aspects and to examine children from 1-9 years for TF. Identified cases of TF were then confirmed and recorded by the certified eye-grader and referred to Community Health Promoters (CHPs) for treatment and follow-up. The questionnaire and the observation checklist were administered through the Kobo Collect smartphone application and data stored in a centralized server which was only accessible by the principal investigator.

## Data analysis

The data was cleaned and analyzed using Stata version 17. Responses were assigned numeric identifiers for quantification, analysis and interpretation. The proportion of children with TF and dirty faces (defined as face with flies, food particles, eye and /or nose secretions) as well as household hygiene practices were descriptively reported. A multivariable logistic regression model using a Generalized Estimated Equations (GEE) was employed to examine the factors that were associated with the prevalence of TF. Domain-specific models (Demographic characteristics, Housing and living conditions, Hygiene and Sanitation Practices, Knowledge on trachoma disease: cause, transmission and prevention), were fitted to identify variables significantly associated with TF prevalence. Statistically significant variables from these models were then included in a final multivariable model to assess their combined effect.

### Results Response rate

At data collection, we were able to achieve a sample of 178 households in Tiaty East Sub County. A response rate of at least 85% in cross sectional studies is considered sufficient to achieve target parameters (23). Within these 178 households, 279 children were observed for TF and facial cleanliness.

### Socio demographic and economic characteristics

Out of the 178 households sampled in this study 45.5% of the caregivers fell under the 21-30 age category, 80.3% were female, 46.1% had some form of education (at least primary school). Additionally, 82% had a form of employment while 43.8% had a monthly household income ranging from KES 1001-4999. Moreover, 31.5% were living in thatched houses and had a mean of 6 household members with 60.1% having one sleeping room. More socio-demographic and socio-economic details of the sample are outlined in *Table 1* and 2.

**Table 1:**
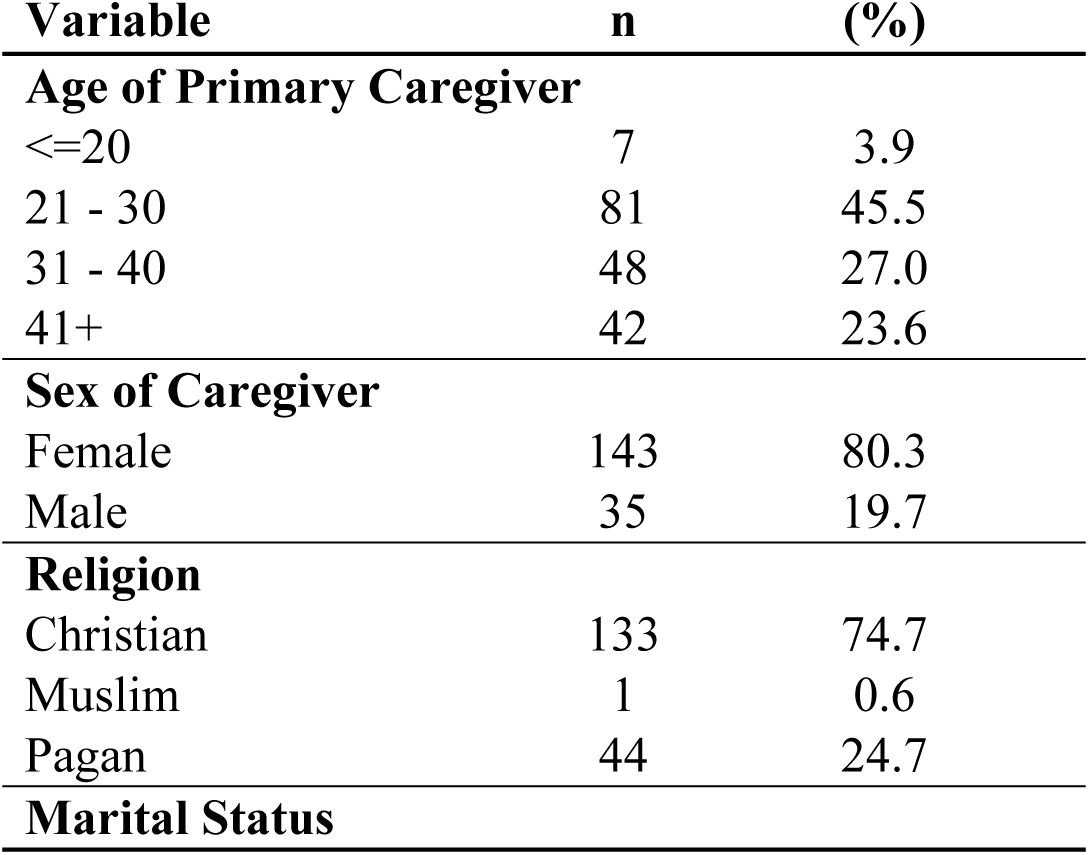
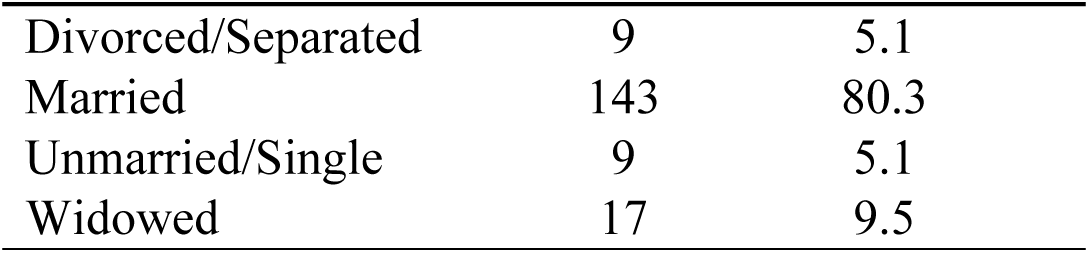
Socio-demographic characteristics.

| Variable | n | (%) |
| --- | --- | --- |
| **Age of Primary Caregiver** |  |  |
| <=20 | 7 | 3.9 |
| 21 - 30 | 81 | 45.5 |
| 31 - 40 | 48 | 27.0 |
| 41+ | 42 | 23.6 |
| **Sex of Caregiver** |  |  |
| Female | 143 | 80.3 |
| Male | 35 | 19.7 |
| **Religion** |  |  |
| Christian | 133 | 74.7 |
| Muslim | 1 | 0.6 |
| Pagan | 44 | 24.7 |
| **Marital Status** |  |  |
| Divorced/Separated | 9 | 5.1 |
| Married | 143 | 80.3 |
| Unmarried/Single | 9 | 5.1 |
| Widowed | 17 | 9.5 |

**Table 2:** Socio-economic characteristics.

| **Variable** | **n** | **(%)** |
| --- | --- | --- |
| **Care giver education level** |  |  |
| Some form of education | 82 | 46.1 |
| No Education | 96 | 53.9 |
| **Employment status of household head** |  |  |
| Employed/Business | 146 | 82.0 |
| Unemployed | 32 | 18.0 |
| **Monthly household Income** |  |  |
| <=1000 | 19 | 10.7 |
| 1001 - 4999 | 78 | 43.8 |
| 5000 - 9999 | 55 | 30.9 |
| 10000+ | 26 | 14.6 |
| **Type of House** |  |  |
| Iron Sheets | 31 | 17.4 |
| Mud | 35 | 19.7 |
| Grass/Thatched | 56 | 31.5 |
| Stone | 27 | 15.1 |
| Timber | 29 | 16.3 |
| **Number of rooms for sleeping** |  |  |
| 1 | 107 | 60.1 |
| 2 | 48 | 27.0 |
| 3 | 18 | 10.1 |
| 4 | 3 | 1.7 |
| 5 | 2 | 1.1 |
| **Number of household members** |  |  |
| Mean: 5.93 (SD ± 2.47), Min 2, Max 17 | | |
| **Means of transport** |  |  |
| None | 162 | 91.0 |
| Motorcycle | 15 | 8.4 |
| Car | 1 | 0.6 |

### Prevalence of trachoma inflammation folliculitis

The study found that 5.02% of 279 children observed in the households had active trachoma. The findings were compared to a previous impact survey report conducted in the same region in 2023, which found a prevalence of 3.02%(17). The difference between the two was not statistically significant (p=0.084) (*Table 3*)

**Table 3:**
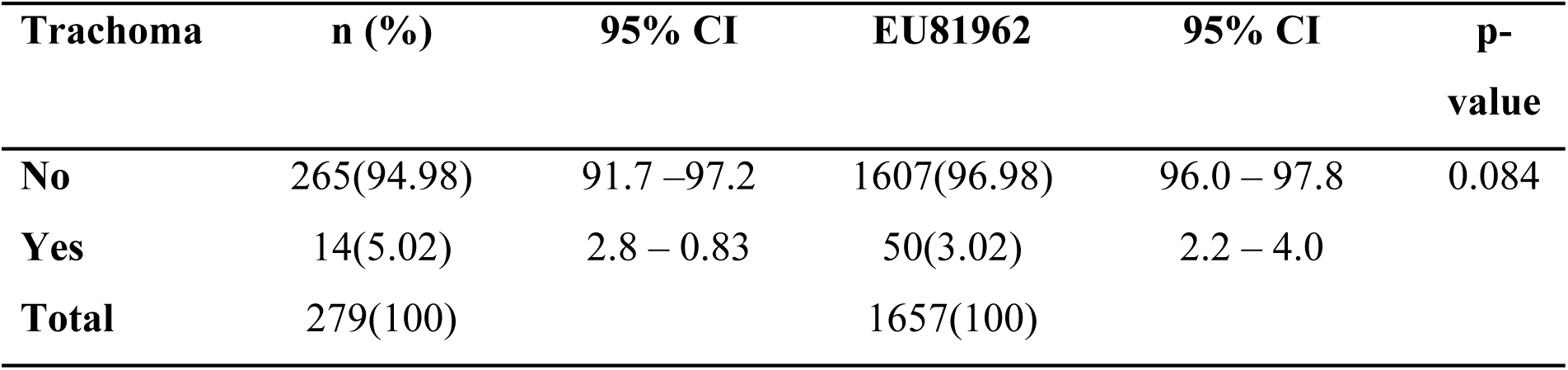
TF Prevalence.

### Facial cleanliness

Facial uncleanliness was observed with 91.8% of children having visibly unclean faces with either dirt on their face, nasal and eye discharge (*Table 4)*

**Table 4:** Facial Cleanliness.

| **Variable** | **n (%)** |
| --- | --- |
| **Any Dirt on the Face** |  |
| No | 188(67.4) |
| Yes | 91(32.6) |
| **Discharge from the eye** |  |
| No | 192(68.8) |
| Yes | 87(31.2) |
| **Discharge from the nose** |  |
| No | 204(73.1) |
| Yes | 75(26.9) |
| **Flies on the face** |  |
| No | 242(86.7) |
| Yes | 37(13.3) |
| **Overall Facial Cleanliness** |  |
| Not Clean | 256(91.8) |
| Clean Face | 23(8.2) |

### Environmental Improvement

#### Hygiene and sanitation facilities

Availability of hygiene and sanitation infrastructure is important for good hygiene practices. Of the sampled households, 78.7% did not have a working latrine in their compound and 94.4% did not have a handwashing station. This shows a gap in some of the key aspects of hygiene and sanitation. Other aspects including household water source, time taken to water source and water storage are shown in *Table 5*

**Table 5:** Household hygiene and sanitation facilities.

| **Variable** | **n (%)** |
| --- | --- |
| **Working latrine in compound** |  |
| No | 140(78.7) |
| Yes | 38(21.3) |
| **Handwashing station** |  |
| No | 168(94.4) |
| Yes | 10(5.6) |
| **Fly Traps** |  |
| No | 161(90.5) |
| Yes | 17(9.5) |
| **Household's primary source of water?** |  |
| Dam | 110(61.8) |
| Piped Water | 13(7.3) |
| Stream/River | 27(15.2) |
| Borehole | 28(15.2) |
| **Time taken to water source (one way)** |  |
| Water source in compound | 5(2.8) |
| Less than 30 minutes | 29(16.3) |
| 30 minutes to 1 hour | 136(76.4) |
| More than 1 hour | 8(4.5) |
| **Water storage** |  |
| Barrels > 20litres | 46(25.8) |
| Containers ≤ 20litres | 123(69.1) |
| Tanks > 1000litres | 9(5.1) |
| **Closed water storage** |  |
| Yes | 121(68.0) |
| No | 57(32.0) |

#### Hygiene and sanitation practices

Following the household hygiene and sanitation facilities reported in *Table 5*, data on hygiene practices show that 94.7% of households that had a functional latrine within the compound did not have all members using it and 99.1%, of households that either did not have functional latrines or did not have all members using the present latrines, defecated in a bush or open compound. All aspects of hygiene and sanitation practices are shown in *Table 6*.

**Table 6:** Household hygiene and sanitation practices.

| Variable | n (%) |
| --- | --- |
| **Face washing frequency** |  |
| Three times a day | 42(23.6) |
| Once a day | 56(31.5) |
| Over three times a day | 25(14.0) |
| Two times a day | 55(30.9) |
| **Face washing means** |  |
| Soap and Water | 86(48.3) |
| Water Only | 92(51.7) |
| **Clothes washing frequency** |  |
| More than once a week | 93(52.2) |
| Once a month | 9(5.1) |
| Once a week | 70(39.3) |
| Two times a month | 6(3.4) |
| **All members using latrine** |  |
| No | 2(5.3) |
| Yes | 36(94.7) |
| **Reason for not using latrine** |  |
| Cultural reasons | 82(57.8) |
| Other Reasons | 2(1.4) |
| Preference | 58(40.8) |
| **Latrine cleaning** |  |
| Daily | 19(50.0) |
| Once a week | 10(26.3) |
| More than a week | 4(10.5) |
| Never | 5(13.6) |
| **If latrine is absent or not using latrine, where they defecate** |  |
| Bush/Compound | 140(99.1) |
| Shared latrines | 2(0.9) |
| **Livestock present** |  |
| Yes | 141(79.2) |
| No | 37(20.8) |
| **Where animals are kept** |  |
| In the house | 21(14.9) |
| 20m or less from the house | 49(48.9) |
| >20m from the house | 51(36.2) |
| **Visible feces on compound** |  |
| No | 57(32.0) |
| Yes | 121(68.0) |
| **Garbage in the compound** |  |
| No | 146(82.0) |
| Yes | 32(18.0) |

### Factors associated with the prevalence of Trachoma Folliculitis

Parents or caregivers above 20 years were significantly less likely to have a child with TF as compared to younger caregivers with less than 20 years. However, the likelihood varied with age, with 21–30 years (AOR = 0.027, 95% CI: 0.002–0.306, *p* = 0.004), 31–40 years (AOR = 0.049, 95% CI: 0.004–0.586, *p* = 0.017), and ≥40 years (AOR = 0.028, 95% CI: 0.002–0.471, *p* = 0.013). Additionally, households with a monthly income of KES 5000-9999 were less likely to have a child with TF (AOR = 0.058, 95% CI: 0.004–0.851, *p* = 0.038), compared to those that earned KES 1000 or less. While households with men as primary care givers were more likely to have a child with TF, the difference was, however, not statistically significant. Caregiver’s marital status, education level and religion were not significantly associated with TF (*Table 7*).

**Table 7:** Association of Demographic characteristics and TF Prevalence.

| Variable | Crudes Odds Ratio | P-value | 95% CI | Adjusted Odds Ratio | P-value | 95% CI |
| --- | --- | --- | --- | --- | --- | --- |
| **Caregiver's Age** |  |  |  |  |  |  |
| $\leq 20$ | REF | | | REF | | |
| $\geq 21$ & $\leq 30$ | 0.042 | $<0.001$ | [0.008, 0.218] | 0.027 | **0.004** | [0.002, 0.306] |
| $>31$ & $\leq 40$ | 0.112 | 0.004 | [0.025, 0.503] | 0.049 | **0.017** | [0.004, 0.586] |
| $\geq 40$ | 0.056 | 0.001 | [0.009, 0.331] | 0.028 | **0.013** | [0.002, 0.471] |
| **Caregiver's Sex** |  |  |  |  |  |  |
| Female | REF |  |  | REF |  |  |
| Male | 3.666 | 0.024 | [1.182, 11.370] | 3.175 | 0.169 | [0.613, 16.461] |
| **Caregiver's Marital Status** |  |  |  |  |  |  |
| Divorced | REF |  |  | REF |  |  |
| Married | 0.366 | 0.346 | [0.045, 2.968] | 0.307 | 0.341 | [0.027, 3.488] |
| Unmarried/Single | 1.751 | 0.643 | [0.164, 18.657] | 0.157 | 0.353 | [0.003, 7.814] |
| Widowed | 0.361 | 0.475 | [0.022, 5.893] | 0.700 | 0.844 | [0.020, 24.543] |
| **Caregiver's Education** |  |  |  |  |  |  |
| No education | REF |  |  | REF |  |  |
| Some level of education | 0.951 | 0.929 | [0.311, 2.909] | 0.892 | 0.898 | [0.155, 5.124] |
| **Household Monthly Income (KES)** |  |  |  |  |  |  |
| $\leq 1000$ | REF | | | REF | | |
| $\geq 1001$ & $\leq 4999$ | 0.458 | 0.264 | [0.116, 1.804] | 0.277 | 0.185 | [0.042, 1.846] |
| >=5000 & <=9999 | 0.079 | 0.043 | [0.007, 0.921] | 0.058 | **0.038** | [0.004, 0.851] |
| >=10000 | 0.184 | 0.150 | [0.018, 1.838] | 0.353 | 0.445 | [0.024, 5.107] |
| **Religion** |  |  |  |  |  |  |
| Christian | REF |  |  | REF |  |  |
| None | 1.941 | 0.247 | [0.631, 5.970] | 4.125 | 0.122 | [0.684, 24.884] |

After adjusting for confounding factors, none of the variables within the domain of Housing and Living Conditions was significantly associated with trachoma prevalence (*Table 8*).

**Table 8:** Association of Housing and Living Conditions and TF Prevalence.

| Variable | Crudes Odds Ratio | P-value | 95% CI | Adjusted Odds Ratio | P-value | 95% CI |
| --- | --- | --- | --- | --- | --- | --- |
| **Type of House** |  |  |  |  |  |  |
| Other (specify) | REF |  |  | REF |  |  |
| Timber/Iron sheet/Stone | 0.494 | 0.254 | [0.147, 1.663] | 0.713 | 0.661 | [0.157, 3.235] |
| **Number of Rooms used for sleeping** |  |  |  |  |  |  |
| 1 room | REF |  |  | REF |  |  |
| 2 rooms | 0.227 | 0.159 | [0.029, 1.782] | 0.264 | 0.250 | [0.027, 2.561] |
| 3 rooms | 0.774 | 0.742 | [0.168, 3.571] | 0.409 | 0.443 | [0.042, 4.014] |
| **Number of Household Members** |  |  |  |  |  |  |
| Less than 4 | REF |  |  | REF |  |  |
| More than 4 | 1.478 | 0.612 | [0.326, 6.704] | 1.374 | 0.701 | [0.271, 6.963] |
| **Livestock kept** |  |  |  |  |  |  |
| No | REF |  |  | *** |  |  |
| Yes | 2.57 | 0.362 | [0.337, 19.584] |  |  |  |
| **Where livestock are kept** |  |  |  |  |  |  |
| In the house | REF |  |  | REF |  |  |
| Outside the house | 0.659 | 0.517 | [0.187, 2.327] | 0.602 | 0.424 | [0.173, 2.093] |

\*\*\*Omitted due to collinearity

After adjusting for confounding factors, the study did not find any significant association between Hygiene and Sanitation Factors, and the prevalence of TF. For instance, washing hands more than once reduced the likelihood of TF infection, but the difference (AOR = 0.790, 95% CI: 0.679– 2.414, *p* = 0.679) compared to those who washed only once a day was not statistically significant. Households that used water and soap to clean their hands and face were less likely to have TF than those that used soap alone even though the difference was not significant (95% CI: 0.240–2.258, *p* = 0.592). Similar results were reported for households that had and used latrines compared to those that did not (AOR = 0.271, 95% CI: 0.062–1.183, *p* = 0.082). The presence of fly traps in the households was also not significantly associated with the prevalence of TF (AOR = 2.267, 95% CI: 0.244–21.048, *p* = 0.472) (*Table 9*).

**Table 9:**
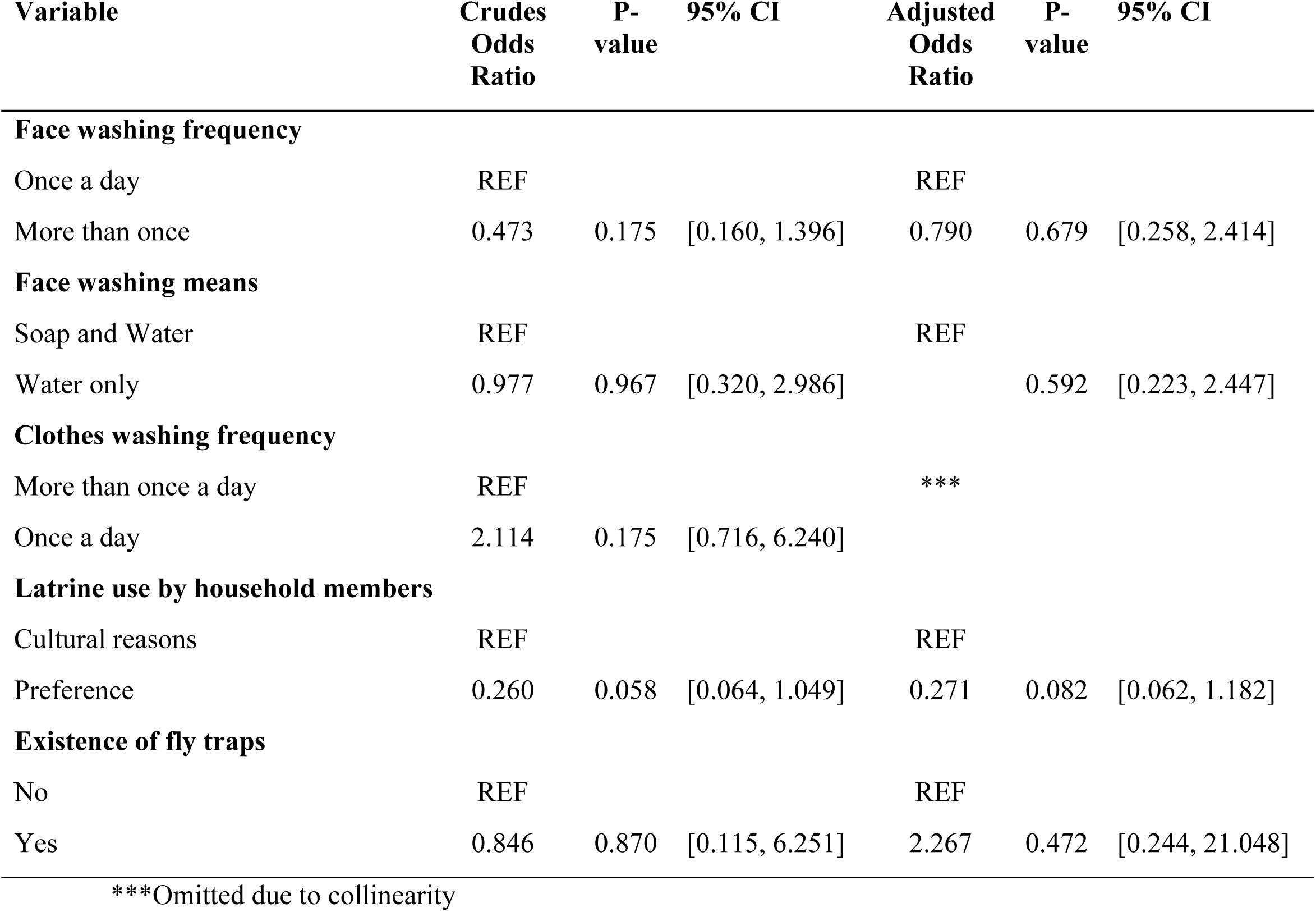
Association of Hygiene and Sanitation Practices and TF Prevalence.

| Variable | Crudes Odds Ratio | P-value | 95% CI | Adjusted Odds Ratio | P-value | 95% CI |
| --- | --- | --- | --- | --- | --- | --- |
| **Face washing frequency** |  |  |  |  |  |  |
| Once a day | REF |  |  | REF |  |  |
| More than once | 0.473 | 0.175 | [0.160, 1.396] | 0.790 | 0.679 | [0.258, 2.414] |
| **Face washing means** |  |  |  |  |  |  |
| Soap and Water | REF |  |  | REF |  |  |
| Water only | 0.977 | 0.967 | [0.320, 2.986] |  | 0.592 | [0.223, 2.447] |
| **Clothes washing frequency** |  |  |  |  |  |  |
| More than once a day | REF |  |  | *** |  |  |
| Once a day | 2.114 | 0.175 | [0.716, 6.240] |  |  |  |
| **Latrine use by household members** |  |  |  |  |  |  |
| Cultural reasons | REF |  |  | REF |  |  |
| Preference | 0.260 | 0.058 | [0.064, 1.049] | 0.271 | 0.082 | [0.062, 1.182] |
| **Existence of fly traps** |  |  |  |  |  |  |
| No | REF |  |  | REF |  |  |
| Yes | 0.846 | 0.870 | [0.115, 6.251] | 2.267 | 0.472 | [0.244, 21.048] |

\*\*\*Omitted due to collinearity

Caregivers’ knowledge of the cause of trachoma was significantly associated with the presence of TF in the household. The households whose care givers believed that trachoma was caused by an infection of the eye and flies were significantly less likely to have a child with TF as compared to those who believed that it was caused by curses or witchcraft (*Table 10*)

**Table 10:** Knowledge on causes of Trachoma and TF Prevalence.

| Variable | Odds Ratio | P-value | 95% CI |
| --- | --- | --- | --- |
| **Cause of Trachoma** |  |  |  |
| Witchcraft/Curse | REF |  |  |
| Flies | 0.14 | **0.034** | 0.023 - 0.860 |
| Infection of the Eye | 0.047 | **0.028** | 0.003 - 0.725 |

Households reporting poor hygiene as a potential cause of trachoma were 8 times more likely to have a child with trachoma compared to those that did not (Adjusted OR = 8.592, 95% CI: 1.133– 65.169, p = 0.037). On the other hand, households where caregivers thought that a dirty environment was associated with trachoma transmission were less likely to have a child with trachoma (Adjusted OR = 0.283, 95% CI: 0.083–0.965, *p* = 0.044). However, knowledge of contact with infected discharge, sharing linen, and presence of flies as possible transmitters were not significantly correlated with TF (*Table 11*).

**Table 11:** Association of Knowledge about Trachoma transmission and TF Prevalence.

| Variable | Crudes Odds Ratio | P-value | 95% CI | Adjusted Odds Ratio | P-value | 95% CI |
| --- | --- | --- | --- | --- | --- | --- |
| **Contact with discharge of infected people** |  |  |  |  |  |  |
| No | REF |  |  |  |  |  |
| Yes | 0.508 | 0.269 | [0.152, 1.690] | 0.459 | 0.173 | [0.150, 1.407] |
| **Flies** |  |  |  |  |  |  |
| No | REF |  |  |  |  |  |
| Yes | 1.379 | 0.693 | [0.280, 6.791] | 1.445 | 0.646 | [0.301, 6.932] |
| **Sharing linen/clothes** |  |  |  |  |  |  |
| No | REF |  |  |  |  |  |
| Yes | 1.017 | 0.978 | [0.315, 3.281] | 0.965 | 0.953 | [0.299, 3.115] |
| **Poor personal hygiene** |  |  |  |  |  |  |
| No | REF |  |  |  |  |  |
| Yes | 4.744 | 0.140 | [0.601, 37.434] | 8.592 | **0.037 \*\*** | [1.133, 65.169] |
| **Dirty environment** |  |  |  |  |  |  |
| Yes | REF |  |  |  |  |  |
| No | 2.804 | 0.101 | [0.817, 9.623] | 0.283 | **0.044 \*\*** | [0.083, 0.965] |

Before adjustment, lack of knowledge regarding preventive measures for trachoma was significantly associated with TF prevalence (Crudes OR = 9.906, 95% CI: 1.675–8.576, *p* = 0.01). There was, however, no significant association between the knowledge of trachoma prevention and presence of TF after adjustment (*Table 12*).

**Table 12:**
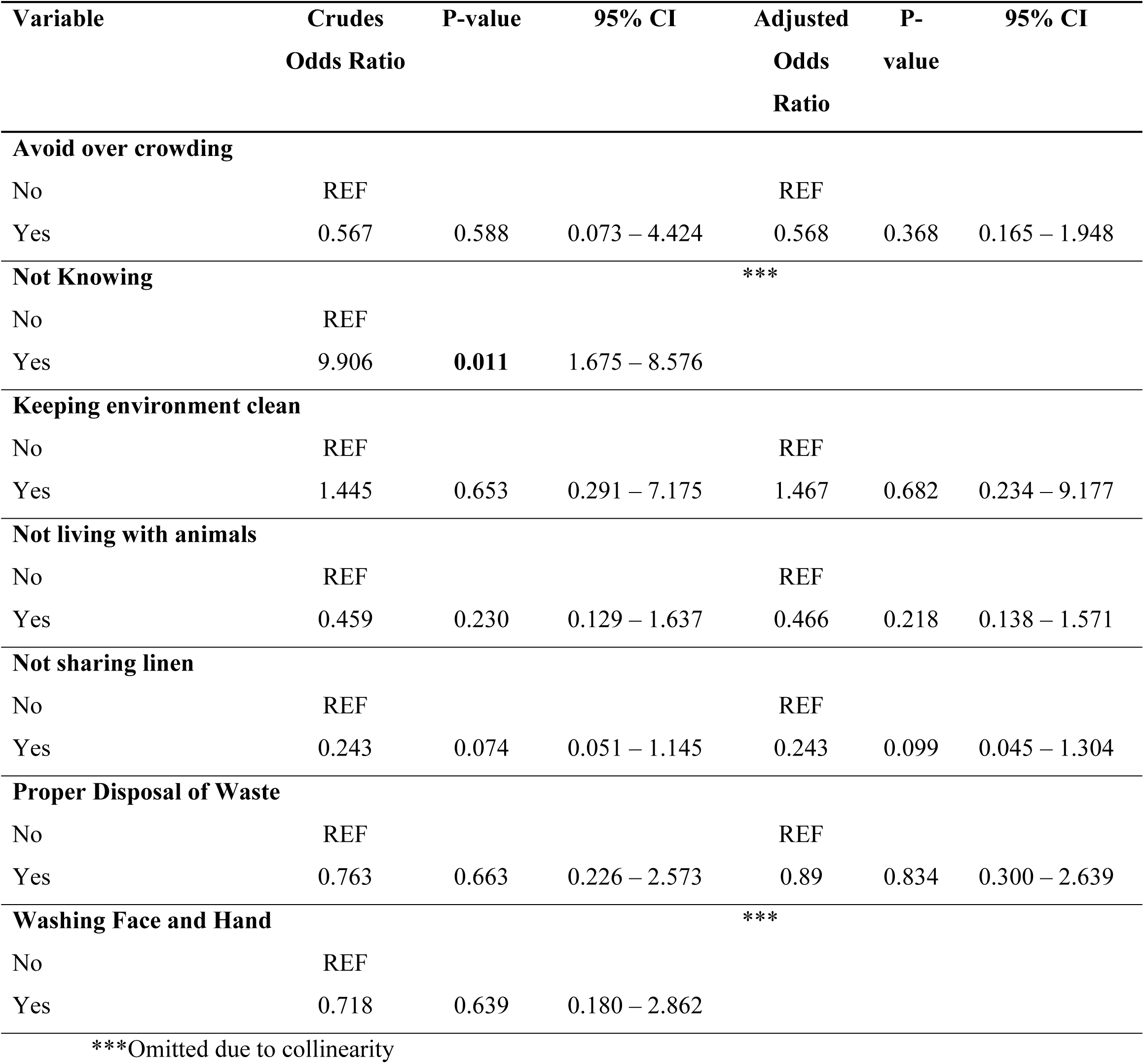
Association of Knowledge on Trachoma prevention and TF Prevalence.

A combined domain-wise multivariate GEE model revealed that the age of the caregiver was significantly correlated with the presence of TF, with older care givers above 20 years being less likely to have a child with trachoma. Knowledge of poor personal hygiene as risk factor was also significantly associated with TF prevalence (AOR = 3.795, p = 0.017). While time taken to fetch water and knowledge of the causes of trachoma were not significantly associated with the disease, they showed a protective trend (*Table 13*).

**Table13:**
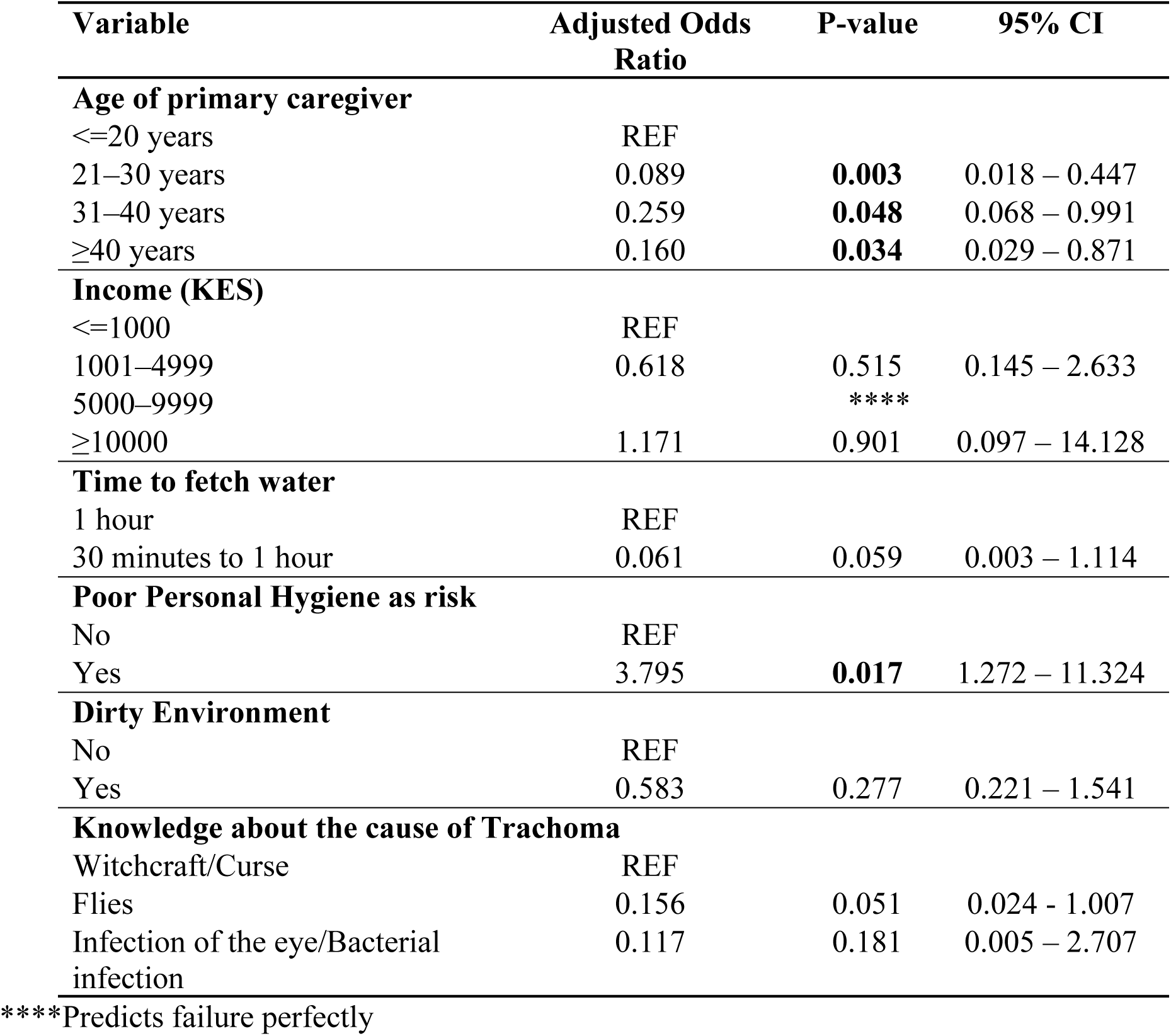
Final model of Factors Associated with TF Prevalence.

## Discussion

### TF prevalence

This study established a trachoma prevalence rate of 5.02% in Tiaty East(a sub EU), which is slightly higher than the WHO trachoma elimination target of <5.00% (4). A post MDA impact survey done in 2023 in Tiaty East and Tiaty West sub counties reported a prevalence of 3.02% (17), which is below the WHO threshold for elimination. In as much as the 2023 impact survey shows a prevalence below the WHO threshold at the EU level, and a decrease from 12.80% in 2018(24), sub EU prevalence reported in our study suggest possible pockets of active trachoma infection in Tiaty East sub county, contributing to the overall EU prevalence. This calls for interventions such as targeted treatment and investment in improving F&E infrastructure and behavior change at the sub-EU level.

### Facial cleanliness

The current study shows facial cleanliness is a challenge in the study area with 91.8% of children observed having dirty faces. In a study conducted in Senegal where the prevalence rates were within the WHO targets, 62.6% of the children had dirty faces (25). In Ethiopia, the country that carries 62% of the global trachoma burden (26), Lakew et al. found that 78.2% of the children had dirty faces(8) which is lower than the current study. Our findings concur with the forementioned studies(8,27), that facial uncleanliness is a persistent issue in some of the most affected African countries. Facial cleanliness remains a critical risk factor for trachoma, as children with ocular or nasal discharge on their faces are significantly more likely to harbor infection, given that these secretions attract eye-seeking flies and facilitate direct transmission through contact(11). This study highlights persistent gaps in facial cleanliness among children in Tiaty East, emphasizing the urgent need for strengthened behavior change communication to address this risk factor.

### Environmental improvement

This study has established that gaps in several environmental improvement infrastructures and practices for trachoma control still exist. On latrine infrastructure and use, our study found that 78.7% of households surveyed lacked latrines, with 99.1% of these practicing open defecation. These findings are consistent with an impact evaluation survey conducted after the last MDA in the region, which reported 82.9% of households without latrines and 82.2% practicing open defecation (17). In contrast, studies from Ethiopia show much higher latrine coverage despite a higher TF prevalence of 21.16% (28). WoldeKidan et al. (2019) found that only 2.3% of households lacked latrines and 0.5% of household members practiced open defecation(29). Such stark differences in latrine ownership and use may be explained by cultural norms and economic contexts. For example, a study in Kenya observed a reluctance among some households to share latrines, even within families(30), highlighting that access alone does not guarantee use. Open defecation sustains fly breeding and facilitates continued transmission of trachoma within communities. These factors pose a challenge in breaking trachoma transmission patterns in the area.

Keeping livestock close to dwellings increases exposure to animal feces, which serve as breeding sites for flies (31). While several studies have shown that domestic animal pens located very close to human dwellings (within the household compound or even inside the house) are associated with a higher risk of active trachoma among children(32–34), 63.8% of our sampled households fell in this category. In addition, 68.0% of compounds had visible feces, reflecting gaps in environmental hygiene similar to those reported in pastoralist communities in Ethiopia(35). Studies from settings with improved sanitation and waste management have documented lower fly densities and reduced trachoma transmission(11). Encouragingly, 82.0% of households reported no garbage within the compound, a positive finding considering that improper solid-waste disposal has been shown to significantly increase trachoma prevalence studies in Ethiopia(36) and other African settings (37). These results highlight both progress and persistent gaps in the environmental improvement component of the SAFE strategy.

Water accessibility is a critical determinant for the effectiveness of the F&E components of the SAFE strategy for trachoma elimination. Beyond availability, it is essential that water sources are safe for household use, as emphasized by WHO(38,39). In the present study, only 22.5% of households reported access to safe water sources (piped water and boreholes), and just 2.8% had these sources located within their compound. Furthermore, for both safe and unsafe water sources, 80.9% of households reported a one-way collection time of 30 minutes or more. These findings highlight a significant gap in accessibility to the very resource necessary for sustaining hygiene and sanitation in a household. Improved access to safe water, within or close to households, not only supports better facial cleanliness and environmental hygiene but also facilitates the broader social and behavioral changes required to interrupt trachoma transmission and reduce the burden of other infectious diseases(8,40).

The presence of hand washing stations in the household or compound is recognized as a key environmental improvement factor for trachoma control, since contaminated hands are a known route of trachoma transmission(41). In our study, 94.4% did not have a hand washing facility. While studies have shown facial cleanliness can also be promoted by presence of handwashing stations which can also be used as face washing stations(40), prevalence of handwashing stations in many trachoma-endemic regions is very low. In Amhara Ethiopia, only 13.9% of households had a hand washing station(42), while a more recent study reported 26.0% having a handwashing facility(43). These consistent findings show a need for more advocacy for hand washing stations that may in turn improve both facial and environmental hygiene aspects for trachoma control.

### Associated factors

Given the relatively poor standards of F&E in Tiaty East Sub-County, as reported in this study, we moved to assess factors associated with the prevalence of TF. The outcome in the different domain fitted models in the present study suggests that, caregivers age, monthly household income, knowledge on causes and some aspects on knowledge of transmission of trachoma were significantly associated with TF in our study area, but only caregivers age and knowledge of poor personal hygiene as risk factors for trachoma were significantly associated with TF prevalence in the final model.

Our study suggests that children with care givers above age 20 years have lower odds of being infected with the blinding disease. While several studies have investigated the factors associated with TF in a similar setting(44–46), few studies noted caregiver age as a possible independent risk factor suggesting a possibility of the predictor being neglected in analysis or not strongly associated as seen in a study in Ethiopia(46). Households earning more were less likely to have a child with TF, a finding that aligns with wider evidence, which indicates that trachoma is highly associated with poverty (47). In addition, while previous studies, found that the size of the family was associated with high levels of transmission (34,48), this study did not find any association between family size and prevalence of TF. The findings in this study also contradict the findings by Kamau (24) and Zambrano et.al., (49) who found a significant negative association between being a Christian and a Muslim and trachoma respectively. This necessitates further research on the topic to establish how various aspects of religion may relate to the prevalence of TF.

While our descriptive statistics show that the study area has major gaps in hygiene and sanitation aspects for trachoma control, we did not find these factors significantly associated with TF in the area. These findings contradict evidence from previous studies, which suggest that poor sanitation and hygiene contribute significantly to the spread of trachoma(50). The study also contradicts the findings by Gebire et al. (2019), whose systematic literature review and meta-analysis established a significant association between absence of latrine, unclean faces of children and no reported use of soap for washing and active trachoma among children. However, the significant association of caregivers knowledge of poor personal hygiene as a risk factor for trachoma emphasizes the findings of several studies that knowledge does not always translate to practice(30,51–53). There is therefore a need to focus on behavioral change in the area and similar endemic settings to break trachoma transmission.

## Conclusion and recommendations

This study suggests the possible presence of pockets of active trachoma in Tiaty East, a trachoma sub-EU. Despite repeated MDA campaigns, the limited household adoption of the F&E components of the SAFE strategy may allow persistence and recrudescence of trachoma. Strengthening F&E remains critical to achieving and sustaining elimination of the disease as a public health importance by the national target year of 2027. Future studies can explore qualitatively the issues leading to poor implementation of F&E in this community to curate context specific interventions.

Although TF prevalence in the area has declined over time through scheduled MDAs in line with WHO protocols, long-term elimination requires a multisectoral approach. The Ministry of Health, Ministry of Water, Sanitation and Hygiene, Ministry of Education, and non-governmental partners should prioritize collaborative programs that expand WASH infrastructure, promote health education, and improve community-level awareness and practices. Such interventions will not only support the sustainable elimination of trachoma and other WASH-related diseases but also enhance the overall quality of life of affected populations.

Ultimately, trachoma elimination is more than a disease control milestone, it is a matter of equity and human rights, preventing avoidable blindness and enabling communities to thrive free from the burden of a preventable disease.

## Limitations of the study

This study highlights important aspects of trachoma control but is subject to certain limitations. First, the data were collected at the sub-EU level, which limits direct comparability with EU-level data. Conducting a similar study in Tiaty West would provide a stronger basis for comparison and help identify which areas carry the greatest burden related to gaps in F&E implementation. Second, the questionnaire relied partly on self-reported information, which may be subject to respondent bias. However, this was mitigated using an observation checklist to validate key responses. Future studies should aim to address these limitations by incorporating broader geographic coverage and triangulating data sources.

## Data Availability

All relevant data are within the manuscript and its Supporting Information files.

https://doi.org/10.5281/zenodo.17260910

## Acknowledgements

We would like to thank everyone who supported this study. This includes the Baringo county and Tiaty East sub county health administration, community health promoters, data collectors and the Pokot communities that voluntarily participated in the study.

Special thanks to the Royal Society of Tropical Medicine and Hygiene (RSTMH) Early Career Grants 2024 through partnership with the Children’s Investments Fund Foundation (CIFF) for funding this study. We are also very grateful to Kenya Medical Research Institute (KEMRI) Graduate School, Eastern and Southern Africa Center for International Parasitic Control (ESACIPAC), Ministry of Health National Neglected Tropical Diseases programme and Jomo Kenyatta University of Agriculture and Technology (JKUAT) School of Public Health for guidance throughout this research. This paper has been published with the permission of the Director General of KEMRI.

## Author contributions

**Conceptualization:** Wanjira Njogu.L, Paul M. Gichuki, George A. Makalliwa,

**Data curation:** Wanjira Njogu.L, Paul M. Gichuki.

**Formal analysis:** Wanjira Njogu.L

**Funding acquisition:** Wanjira Njogu.L, Paul M. Gichuki.

**Investigation:** Wanjira Njogu.L, George A. Makalliwa, Bridget Kimani, Kellen Karimi, Zephania Kibet, Doris Njomo, Titus Watitu, Paul M. Gichuki.

**Methodology:** Wanjira Njogu.L, George A. Makalliwa, Bridget Kimani, Kellen Karimi, Zephania Kibet, Doris Njomo, Titus Watitu, Paul M. Gichuki.

**Project administration:** Bridget Kimani, Kellen Karimi, Zephania Kibet, Titus Watitu

**Resources:** Wanjira Njogu.L, Paul M. Gichuki.

**Software:** Wanjira Njogu.L

**Supervision:** George A. Makalliwa, Kellen Karimi, Doris Njomo, Titus Watitu, Paul M. Gichuki.

**Validation:** Kellen Karimi, Doris Njomo, Titus Watitu, Paul M. Gichuki.

**Visualization:** Wanjira Njogu.L

**Writing-original draft:** Wanjira Njogu.L,

**Writing-Review and editing:** Wanjira Njogu.L, George A. Makalliwa, Bridget Kimani, Kellen Karimi, Zephania Kibet, Doris Njomo, Titus Watitu, Paul M. Gichuki.

